# Researchers’ Behavioral Intentions to Engage in Knowledge Translation Activities and Their Association with Bibliometric Indicators: A Theory of Planned Behavior-Informed Questionnaire Study

**DOI:** 10.64898/2026.10.12.26365363

**Authors:** Ayat Ahmadi, Mina Ghourbanpour, Hanieh Sadat Sajadi, Hamid Esmaeilzadeh, Samaneh Orouji, Maryam Zahmatkesh, Azadeh Sayarifard

**Author notes:** **Corresponding author:** Dr. Azadeh Sayarifard, Deputy Director, Research and Development Center, Tehran University of Medical Sciences.

## Abstract

**Background:** Knowledge translation (KT) is essential for ensuring that research evidence informs policy and practice. However, little is known about researchers’ intentions to engage in KT activities or their relationship with conventional bibliometric indicators. This study aimed to (1) assess researchers’ intentions to engage in KT activities using a questionnaire informed by the Theory of Planned Behavior (TPB), and (2) examine the association between these intentions and individual bibliometric indicators.

**Methods:** A cross-sectional survey was conducted among faculty members at Tehran University of Medical Sciences. A questionnaire informed by selected constructs of the TPB was developed to assess researchers’ intentions to engage in activities across the seven components the action cycle of Knowledge-to-Action (KTA) framework. The instrument was validated through several rounds of expert review and then with a sample of potential participants. Bibliometric indicators (H-index, Number of Publications, Number of Citations, First-Author Publications, Corresponding Authorship, International Publications and Self-Citations) were extracted from the national academic registry. To assess representativeness, bibliometric characteristics of respondents were compared with those of a random sample of non-respondents drawn from the same registry. Confirmatory Factor Analysis (CFA) validated the intention construct, and Structural Equation Modeling examined associations with demographic and bibliometric variables.

**Results:** A total of 143 researchers participated. Intention scores were highest for adapting knowledge to local contexts and identifying barriers and facilitators, and lowest for monitoring and sustaining knowledge use. CFA supported a unidimensional intention construct with favorable model-fit indicators. No bibliometric indicator was significantly associated with KT intention. Age was the only significant predictor. Respondents had stronger bibliometric profiles than non-respondents, indicating potential non-response bias.

**Conclusions:** Researchers reported relatively strong intentions to engage in KT, but these intentions were not associated with conventional bibliometric indicators. Age was the only significant predictor. Overall, findings suggest that traditional academic metrics may not reflect KT intentions and highlight the need to strengthen institutional incentives for KT.

## Introduction

Knowledge translation is a dynamic and iterative process that aims to bridge the gap between research evidence and its application in health systems. By facilitating the uptake of scientific findings into policy and practice, KT enhances the societal value of research and contributes to improved health outcomes, more efficient use of resources, and evidence-informed decision making. KT encompasses a broad range of activities, including stakeholder engagement, priority setting, knowledge synthesis, dissemination, implementation, and evaluation, all of which require ongoing interaction between researchers and knowledge users (1, 2).

Despite its recognized importance, the integration of KT into routine academic practice remains challenging. Researchers face numerous barriers to engaging in KT activities, including limited time, insufficient organizational support, lack of KT-related competencies, and academic cultures that prioritize traditional scholarly outputs over societal impact (3, 4). Moreover, the absence of standardized approaches for evaluating KT effectiveness limits its visibility within academic performance systems (5). Consequently, many KT activities remain underutilized or inconsistently implemented despite growing expectations for research to demonstrate real-world impact (6-8).

Addressing these challenges requires attention not only to structural barriers but also to the behavioral determinants of researchers’ engagement in KT activities. Since KT activities are largely discretionary and often extend beyond conventional research responsibilities, their successful implementation depends substantially on researchers’ willingness and motivation to undertake them. Behavioral theories suggest that intention is a critical precursor to action and an important target for intervention when seeking to promote behavior change. According to the Theory of Planned Behavior (TPB), intention is the most proximal determinant of behavior and reflects an individual’s readiness to perform a given action. Intention is shaped by attitudes toward the behavior, perceived social expectations, and perceived behavioral control, and has consistently been shown to predict a wide range of professional and organizational behaviors (9, 10). Assessing researchers’ intentions to engage in KT activities can therefore provide valuable insight into the likelihood of future engagement and help determine whether interventions are needed to strengthen organizational readiness for KT (11).

Beyond individual motivations, researchers’ intentions may also be influenced by the incentive structures within which they work. Academic evaluation and promotion systems signal which activities are valued and rewarded, thereby shaping researchers’ priorities and behavioral choices. However, in many universities and research institutions, performance assessment continues to rely predominantly on bibliometric indicators such as publication counts, citation metrics, and the h-index (12, 13). While these measures are useful for assessing scientific productivity, they have been criticized for inadequately capturing societal impact, stakeholder engagement, and KT-related activities. In Iran, bibliometric indicators remain central components of academic evaluation systems (14), despite increasing national emphasis on knowledge translation and research impact (15).

Whether current evaluation systems are aligned with researchers’ motivation to engage in KT remains unclear. If researchers with stronger intentions toward KT also perform well according to conventional bibliometric indicators, this may suggest a degree of compatibility between existing incentive systems and KT goals. Conversely, the absence of such a relationship may indicate that prevailing evaluation mechanisms neither encourage nor reward activities that promote research utilization and societal impact.

There is limited evidence on researchers’ intentions to engage in KT activities using theory-based approaches. Furthermore, no study in Iran has examined whether such intentions are associated with commonly used bibliometric indicators. Addressing this gap is important both for understanding the current level of readiness for KT among researchers and for assessing the extent to which existing academic incentive systems are aligned with KT objectives.

Therefore, this study aimed to (1) assess researchers’ intentions to engage in KT activities using a questionnaire informed by the TPB, and (2) examine the association between KT intention and individual bibliometric indicators commonly used in academic performance evaluation.

## Methods

### Study Design and Participants

A cross-sectional study was conducted among faculty members at Tehran University of Medical Sciences (TUMS). An institutional list of 1,940 faculty members, including email addresses and available phone numbers, was obtained from the university. All faculty members were invited to participate via an electronic questionnaire, followed by one reminder email and a subsequent mobile text message to non-respondents.

### Development of the Data Collection Tool

An electronic data collection tool was developed by integrating the Knowledge-to-Action (KTA) framework and the Theory of Planned Behavior (TPB). The KTA framework provided the conceptual structure of the questionnaire through its seven action cycle components, while the TPB informed the development of questionnaire items by capturing the behavioral determinants of researchers’ engagement in each component. The KTA framework distinguishes between knowledge creation and the action cycle, with the latter emphasizing the collaborative and iterative processes required to adapt, implement, monitor, and sustain knowledge use in real-world settings (2, 16). Given that successful engagement in these action cycle components is influenced by researchers’ motivation and behavioral intention (11), questionnaire items were developed following TPB questionnaire development guidelines (17). The initial questionnaire was reviewed by an expert panel in several rounds. Following the necessary revisions, the final questionnaire was examined by 12 potential participants to assess its applicability using the think-aloud method (18).

The final tool included 10 items measuring intention to engage in the seven components of the Action Cycle of the KTA model. Demographic and career-related variables—including age, years since graduation, gender, academic rank, and bibliometric characteristics retrieved from the national registry of academic research activities (www.ISID.ac.ir)—were also collected. Bibliometric indicators included h-index, total publications, citation, first-author and corresponding-author publications, international collaborations (i.e., publications with at least one co-author affiliated with an institution outside Iran), and self-citations. Behavioral intention was conceptualized as a latent construct representing researchers’ motivation to engage in KT activities across the seven components of the KTA Action Cycle. The 10 intention items were scored on a 5-point Likert scale (1= strongly disagree to 5 = strongly agree) (Supplemental File 1).

### Data Collection Procedures

The online questionnaire link was distributed to all faculty members in May 2025. After survey completion, bibliometric data for respondents were extracted from their public profiles in the national registry and linked to survey responses. All personal identifiers were then permanently removed, and analyses were conducted on de-identified data.

### Statistical Analysis

The 10 intention items were used to extract the latent behavioral intention construct based on their shared variance. Structural Equation Modeling (SEM) was applied to estimate the simultaneous direct effects of demographic and bibliometric variables on the behavioral intention construct. Standardized coefficients were reported to facilitate comparison across predictors. Model fit was evaluated using standard indices, including RMSEA, χ^2^/df, GFI, and PCFI. Statistically suggested correlations between error terms were incorporated when conceptually justified and when they improved model fit. To evaluate representativeness, bibliometric indicators of respondents were compared with those of a random sample of 65 non-respondents using the same national registry data.

### Ethical Considerations

The study was approved by the Tehran University of Medical Sciences Ethics Committee (approval code: IR.TUMS.NIHR.REC.1401.024). Electronic informed consent was obtained from all participants prior to data collection.

## Results

A total of 143 faculty members participated in the study (response rate: 7.3%). The mean age of respondents was 51.6 years (95% CI: 50.1–53.0), with the largest proportion aged 50–60 years (41.7%), followed by 40–50 years (34.5%). Females comprised 46.2% of the sample and were more represented among participants under 40 years of age, whereas males predominated among those over 60.

Bibliometric indicators showed substantial variability. The median number of publications was 53 (IQR: 28–115.8), and the median citation was 619 (IQR: 236–2426). Respondents had a median h-index of 13 (IQR: 8–24) and a median of 9 international publications (IQR: 3.5–28). Table 1 summarizes participants’ demographic and bibliometric characteristics.

**Table 1.** Baseline characteristics and bibliometric indicators of participants (N=143)

| Variable | Descriptive Statistics |
| --- | --- |
| Gender (Female) [n (%)] | 66(46.15) |
| Age Category (years) [n (%)] |  |
| $\leq 40$ | 16(11.51) |
| 40–50 | 48(34.53) |
| 50–60 | 58(41.73) |
| $> 60$ | 17(12.23) |
| Years Since Graduation [Median (IQR)] | 14(9 – 21) |
| Number of Publications [Median (IQR)] | 53(28 – 115.75) |
| H-index [Median (IQR)] | 13(8 – 24) |
| Number of Citations [Median (IQR)] | 619 (236 – 2426) |
| First-author Publications [Median (IQR)] | 9 (5 – 16) |
| Corresponding-author Publications [Median (IQR)] | 13 (4 – 29) |
| International Publications [Median (IQR)] | 9 (3.5 – 28) |
| Self-citations [Median (IQR)] | 3 (1 – 7) |

Among the ten evaluated items covering the seven components of the KTA Cycle, the highest mean intention scores were observed for “Adopt Knowledge to Local Context” (Mean = 4.20, SE = 0.09) and the second item of “Assess Barriers and Facilitators to Knowledge Use” (Mean = 3.94, SE = 0.10). The lowest mean intention scores were related to “Sustain Knowledge Use” (Mean = 3.13, SE = 0.11) and “Monitor Knowledge Use” (Mean = 3.18, SE = 0.10). Item-level means and factor loadings are presented in Table 2.

**Table 2.** Factor loadings of observed indicators of the Action cycle in Knowledge-to-Action framework (KTA)

| Component of Action Cycle in KTA framework | Measured item for intention | Mean (Range: 1-5) | Factor Loading (Un standardized) | SE | P |
| --- | --- | --- | --- | --- | --- |
| Identify Problem | Refining the research questions in collaboration of research users | 3.35 | 0.66 | 0.10 | <0.01 |
|  | Addressing research questions initiated by research users | 3.49 | 0.42 | 0.11 | <0.01 |
| Adopt Knowledge to Local Context | Perform research in a way that findings are adaptable to local context | 4.20 | 0.64 | 0.09 | <0.01 |
| Assess Barriers and Facilitators to Knowledge Use | Explaining benefits of using research findings to users | 3.48 | 0.77 | 0.11 | <0.01 |
|  | Gather information on barriers and facilitators of implementing research findings | 3.94 | 0.79 | 0.10 | <0.01 |
| Select, Tilor and Implement Interventions | Plan appropriate strategies to address barriers and facilitators | 3.53 | 0.72 | 0.11 | <0.01 |
|  | Applying effective strategies, beyond publication, to communicate research findings | 3.47 | 0.84 | 0.10 | <0.01 |
| Monitor Knowledge Use | Gathering data on whether users are informed about findings | 3.18 | 0.81 | 0.10 | <0.01 |
| Evaluate Outcomes | Gathering data on effectiveness of communication/implementation strategy | 3.20 | 0.69 | 0.10 | <0.01 |
| Sustain Knowledge Use | Gathering data on sustainable implementation of findings | 3.13 | 0.64 | 0.11 | <0.01 |

RMSEA: 0.049, CMIN/DF= 1.34, GFI=0.944, PCFI=0.679

In the confirmatory factor analysis, all ten items loaded significantly onto the latent intention factor (p < 0.01). The highest standardized loading was observed for “Select, Tailor, and Implement Interventions” (loading = 0.84, p < 0.01), and the lowest—yet statistically significant—loading was for “Identify Problem” (loading = 0.42, p < 0.01). Model fit indices indicated good fit: RMSEA = 0.05, χ^2^/df = 1.34, GFI = 0.94, PCFI = 0.67. Four conceptually justified error-term correlations were added to improve model fit. Standardized loadings are shown in Figure 1.

**Figure 1.**
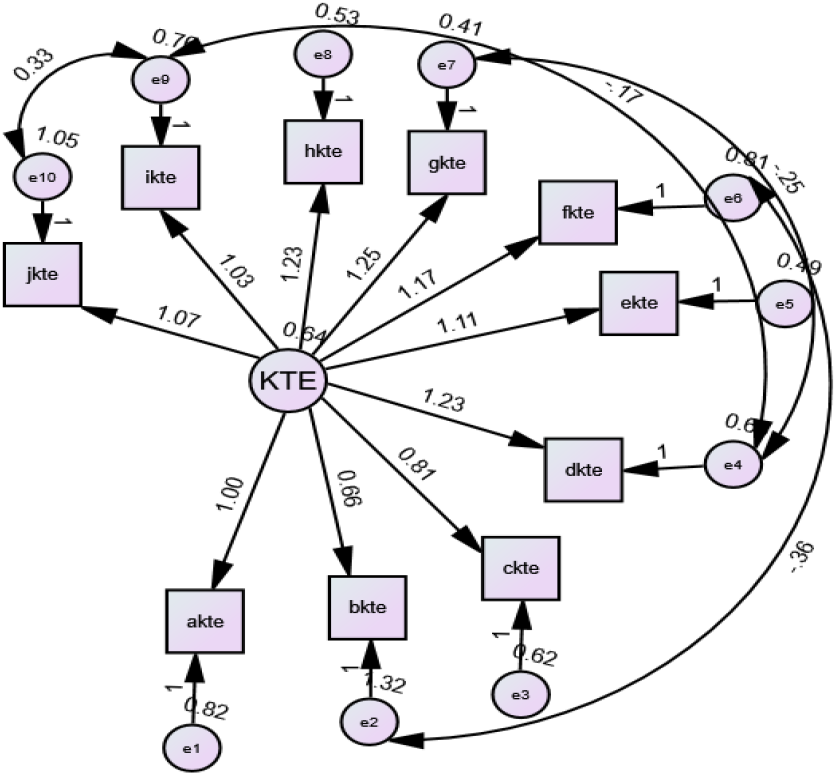
Standardized factor loadings of 10 items measuring the behavioral intention construct across the seven components of the Action Cycle in the Knowledge-to-Action (KTA) framework. RMSEA: 0.049, CMIN/DF= 1.34, GFI=0.944, PCFI=0.679 KTE: Behavioral intention to undertake components of the Action Cycle in Knowledge-to-Action (KTA) framework akte: refining the research questions in collaboration of research users (Component of the Action Cycle in KTA: Identify Problem) bkte: addressing research questions initiated by research users (Component of the Action Cycle in KTA: Identify Problem) ckte: perform research in a way that findings are adaptable to local context (Component of the Action Cycle in KTA: Adopt Knowledge to Local Context) dkte: explain benefits of using research findings to users (Component of the Action Cycle in KTA: Assess Barriers and Facilitators to Knowledge Use) ekte: gather information on barriers and facilitators of implementing research findings (Component of the Action Cycle in KTA: Assess Barriers and Facilitators to Knowledge Use) fkte: plan appropriate strategies to address barriers and facilitators (Component of the Action Cycle in KTA: Select, Tilor and Implement Interventions) gkte: applying effective strategies, beyond publication, to communicate research findings (Component of the Action Cycle in KTA: Select, Tilor and Implement Interventions) hkte: gather data on whether users are informed about findings (Component of the Action Cycle in KTA: Monitor Knowledge Use) ikte: gather data on effectiveness of communication/implementation strategy (Component of the Action Cycle in KTA: Evaluate Outcomes) jkte: gather data on sustainable implementation of findings (Component of the Action Cycle in KTA: Sustain Knowledge Use)

SEM examined the direct effects of demographic and bibliometric variables on the behavioral intention construct. Age was the only statistically significant predictor (standardized coefficient = 0.20, p = 0.02)(Figure 2), indicating that older researchers reported stronger intentions to engage in KT activities.

**Figure 2.**
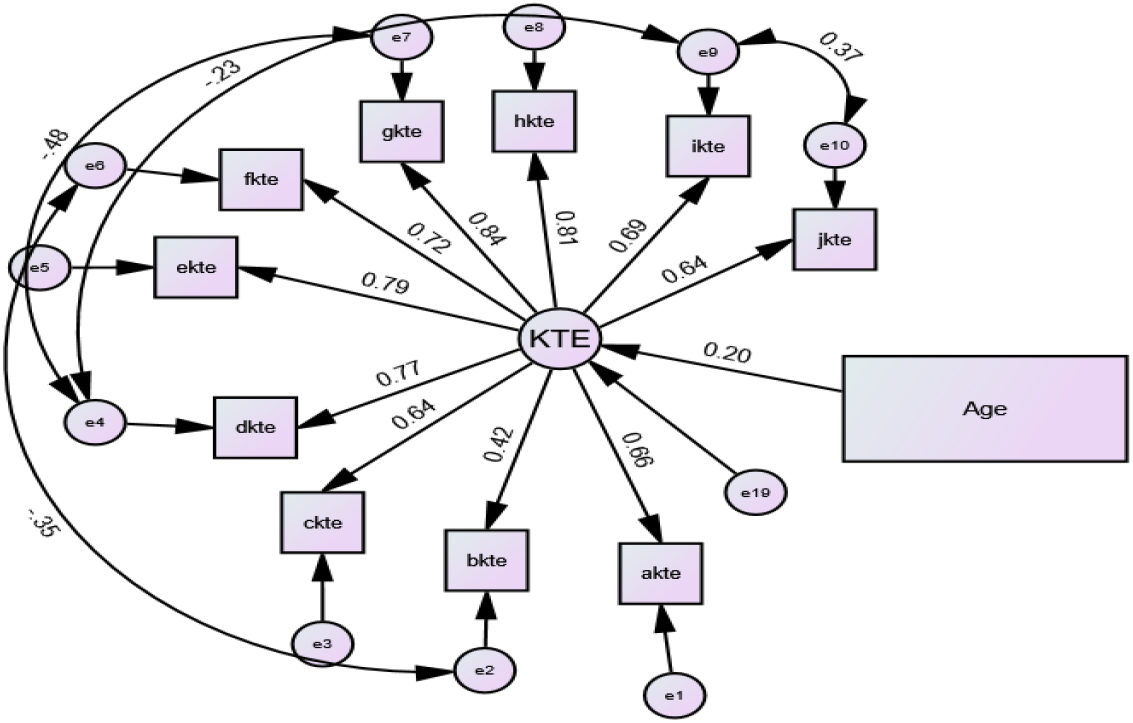
Standardized direct effects of age on behavioral intention to engage in the seven components of the Action Cycle in the Knowledge-to-Action (KTA) framework, as measured by 10 items. KTE: Behavioral intention to undertake components of the Action Cycle in Knowledge-to-Action (KTA) framework akte: refining the research questions in collaboration of research users (Component of the Action Cycle in KTA: Identify Problem) bkte: addressing research questions initiated by research users (Component of the Action Cycle in KTA: Identify Problem) ckte: perform research in a way that findings are adaptable to local context (Component of the Action Cycle in KTA: Adopt Knowledge to Local Context) dkte: explain benefits of using research findings to users (Component of the Action Cycle in KTA: Assess Barriers and Facilitators to Knowledge Use) ekte: gather information on barriers and facilitators of implementing research findings (Component of the Action Cycle in KTA: Assess Barriers and Facilitators to Knowledge Use) fkte: plan appropriate strategies to address barriers and facilitators (Component of the Action Cycle in KTA: Select, Tilor and Implement Interventions) gkte: applying effective strategies, beyond publication, to communicate research findings (Component of the Action Cycle in KTA: Select, Tilor and Implement Interventions) hkte: gather data on whether users are informed about findings (Component of the Action Cycle in KTA: Monitor Knowledge Use) ikte: gather data on effectiveness of communication/implementation strategy (Component of the Action Cycle in KTA: Evaluate Outcomes) jkte: gather data on sustainable implementation of findings (Component of the Action Cycle in KTA: Sustain Knowledge Use)

No significant associations were found between KT intention and other demographic variables or bibliometric indicators. Table 3 presents the standardized effects of all predictors.

**Table 3.**
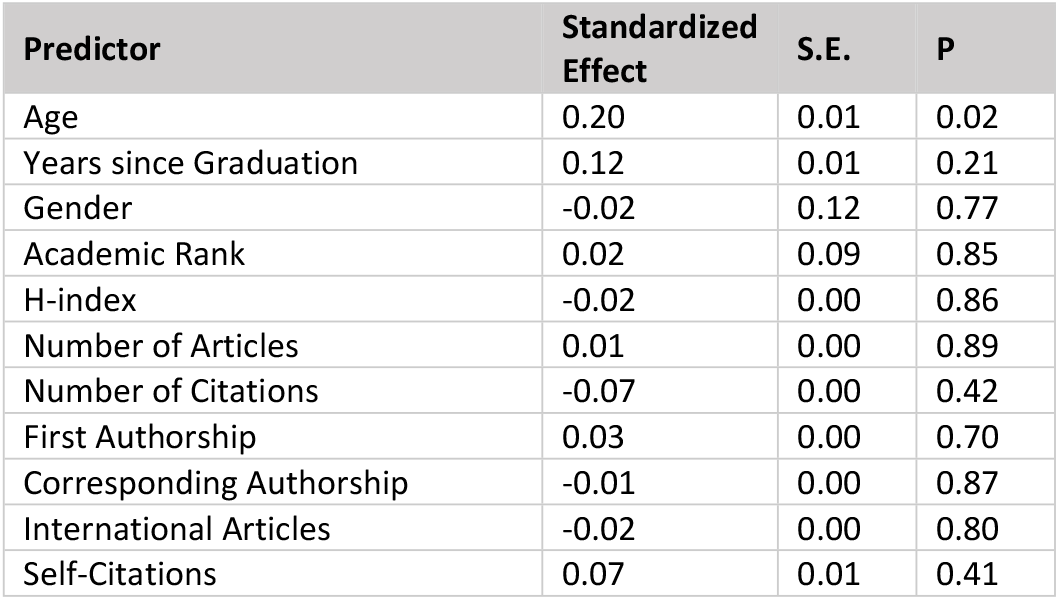
Standardized direct effects of demographic and bibliometric variables on the behavioral intention to engage in the seven components of the Action Cycle in the Knowledge-to-Action (KTA) framework.

| Predictor | Standardized Effect | S.E. | P |
| --- | --- | --- | --- |
| Age | 0.20 | 0.01 | 0.02 |
| Years since Graduation | 0.12 | 0.01 | 0.21 |
| Gender | -0.02 | 0.12 | 0.77 |
| Academic Rank | 0.02 | 0.09 | 0.85 |
| H-index | -0.02 | 0.00 | 0.86 |
| Number of Articles | 0.01 | 0.00 | 0.89 |
| Number of Citations | -0.07 | 0.00 | 0.42 |
| First Authorship | 0.03 | 0.00 | 0.70 |
| Corresponding Authorship | -0.01 | 0.00 | 0.87 |
| International Articles | -0.02 | 0.00 | 0.80 |
| Self-Citations | 0.07 | 0.01 | 0.41 |

To assess representativeness, bibliometric indicators of respondents were compared with those of a random sample of 65 non-respondents. Respondents had significantly higher values for h-index (median 13 vs. 9; p < 0.01), number of publications (median 53 vs. 35; p = 0.02), and citation counts (median 619 vs. 319; p < 0.01). Comparative results are shown in Table 4.

**Table 4.** Comparison of bibliometric indicators between responders and non-responders.

| Variable | Responder(n=143) Median [IQR] | Non-responder (n= 65) Median [IQR] | p-value |
| --- | --- | --- | --- |
| H-index | 13[8 - 24] | 9[4 – 16] | <0.01 |
| Number of Publications | 53[28 - 115.75] | 35 [12 – 60] | 0.02 |
| Number of Citations | 619 [236 – 2426] | 319 [82 – 882] | <0.01 |
| First-Author Publications | 9 [5 – 16] | 5 [3 – 12] | 0.01 |
| Corresponding Authorship | 13 [4 – 29] | 7 [2 – 16] | <0.01 |
| International Publications | 9 [3.5 – 28] | 5 [1 – 11] | <0.01 |
| Self-Citations | 3 [1 – 7] | 2 [ 0.6 – 4] | 0.08 |

## Discussion

This study investigated researchers’ behavioral intentions to engage in KT activities and examined whether these intentions were associated with individual bibliometric indicators. Overall, participants reported relatively strong intentions to engage in KT activities, particularly those related to adapting knowledge to local contexts and assessing barriers and facilitators to knowledge use. In contrast, intentions were weakest for activities requiring sustained engagement, such as monitoring knowledge use and supporting its long-term sustainability. No bibliometric indicator was significantly associated with KT intention. These findings suggest that although researchers express a strong willingness to participate in KT activities, this intention is not reflected in conventional measures of academic productivity, highlighting that KT engagement may be driven by factors beyond bibliometric performance.

A key finding of this study was that researchers demonstrated relatively strong intentions to engage in KT activities, indicating a generally positive willingness to bridge the “know-do” gap. This finding aligns with previous studies reporting that health researchers increasingly view KT as a professional duty and a fundamental requirement of their work (19). Specifically, participants expressed the strongest intentions for activities related to adapting knowledge to local contexts and assessing barriers and facilitators. These early-stage activities are often perceived as more congruent with traditional academic roles and expertise. The literature underscores that such preliminary work is vital; for instance, a deep understanding of local context and the identification of social or structural stigmas are essential prerequisites for the successful adoption of evidence-based practices (20).

In contrast, intentions were weakest for activities involving monitoring knowledge use and supporting long-term sustainability. This pattern likely reflects the broader challenges of maintaining KT processes, which require ongoing collaboration and sustained institutional support. Our findings echo an observed “evaluation gap” in the field, where researchers often omit monitoring and evaluation from their mental models of KT, sometimes viewing these tasks as falling outside the scope of “research” (19). Furthermore, the complexity of the policymaking process is frequently underestimated; researchers may naively dismiss KT as a straightforward “technocratic exercise” rather than a nuanced political process (21). Successfully navigating these later stages requires specific “soft skills”—such as networking and emotional intelligence—that are often absent from traditional academic training (21). Consequently, researchers across all levels of experience continue to report significant struggles in engaging effectively with stakeholders and policy makers to optimize the real-world impact of their findings (22).

Another key finding of this study was the absence of a significant association between researchers’ intentions to engage in KT activities and conventional bibliometric indicators, including publication output, citation, h-index, authorship patterns, and international collaborations. This finding suggests that traditional indicators of academic productivity do not necessarily reflect researchers’ willingness to engage in KT activities. It also reinforces concerns that research evaluation systems relying predominantly on bibliometric indicators may overlook behaviors that are essential for achieving research impact beyond academia (13, 23).

One possible explanation is that effective KT extends well beyond the production and dissemination of scientific publications. It requires sustained stakeholder engagement, adaptation of evidence to local contexts, continuous monitoring of knowledge use, and long-term implementation support. These activities are resource-intensive, collaborative, and dependent on a broad range of interpersonal, organizational, and communication skills (3, 24). Consequently, when promotion and evaluation systems primarily reward publication and citation metrics, researchers may have fewer incentives to invest time and effort in these demanding KT activities (13, 25).

Our findings therefore lend support to ongoing calls for more comprehensive approaches to research assessment. Aligning health research evaluation systems with the principles of responsible research assessment may encourage greater engagement in KT by recognizing activities that contribute to societal impact in addition to traditional academic outputs. Such changes may also strengthen the subjective norms and perceived behavioral control that influence KT intention and facilitate the translation of intention into actual KT behavior (26-28).

Several international initiatives have advocated for this broader perspective on research assessment. For example, the UK Research Excellence Framework (A) evaluates research through a combination of scientific quality, research environment, and demonstrable societal impact rather than relying solely on bibliometric measures (29). Although the REF has been criticized for its administrative burden and potential unintended consequences, it has helped establish research impact as a core objective of universities and has encouraged greater institutional attention to KT activities (30, 31). Likewise, the San Francisco Declaration on Research Assessment (DORA) discourages the use of journal-based metrics as proxies for research quality (32), while the Hong Kong Principles advocate evaluating researchers according to responsible research practices, transparency, rigor, and societal contributions alongside conventional scholarly outputs (33). Collectively, these initiatives underscore the need for evaluation systems that recognize and reward the diverse activities required to achieve meaningful real-world impact.

In our study age emerged as the only significant predictor of KT intention, with older researchers reporting stronger motivation to engage in KT activities. This may reflect differences in career priorities: early-career researchers often face pressure to build publication records and secure promotion, whereas senior researchers may have greater autonomy, established networks, and stronger motivation to pursue activities that enhance real-world impact (34, 35). Although academic rank was not significantly associated with intention, the observed age effect suggests that KT motivation may vary across career stages, consistent with previous work on researcher incentives and professional identity (13).

This study also contributes methodologically by applying TPB guidelines to operationalize KT intention across the full action cycle of the KTA framework (2, 9). We utilized established TPB construction guidelines to develop the data collection tool, which enhanced construct validity and strengthened the measurement of participants’ intentions (17). The use of CFA and SEM enabled rigorous validation of the latent intention construct and simultaneous estimation of predictor effects (36). The acceptable model-fit indicators support the validity of this approach and demonstrate the feasibility of measuring KT intention comprehensively rather than focusing on isolated KT behaviors.

The study’s low response rate raises concern about representativeness and sample size. We compared the characteristics of a random sample of non-respondents with those of respondents to assess the potential impact of non-response bias. Respondents had stronger bibliometric profiles than non-respondents, suggesting that the sample may overrepresented more research-active faculty members. It is also plausible that individuals with greater interest in KT or research impact were more likely to participate. This self-selection may have reduced variability in bibliometric indicators and limited the ability to detect associations. Future studies could consider strategies such as inverse probability weighting or targeted sampling to mitigate non-response bias and enhance generalizability (37).

Despite these limitations, the findings provide important insights into researchers’ intentions to engage in KT within academic settings. The absence of an association between KT intention and conventional bibliometric indicators suggests that traditional measures of academic performance may not adequately reflect researchers’ readiness to engage in KT activities. These findings support ongoing efforts to align institutional evaluation systems with broader indicators of research impact and responsible research assessment, thereby creating environments that better support the integration of research into policy and practice.

## Conclusion

Although academic researchers in this study reported relatively strong intentions to engage in KT activities, these intentions were not significantly associated with conventional bibliometric indicators, including publication output, citation metrics, or h-index. Age was the only significant predictor of KT intention, suggesting that researchers’ willingness to engage in KT may vary across career stages. Together, these findings indicate that traditional measures of academic performance may not adequately reflect researchers’ intentions to undertake KT activities. Beyond the study setting, they provide empirical support for ongoing efforts to reform research assessment in academic systems that rely predominantly on bibliometric indicators, particularly in low- and middle-income countries seeking to strengthen the societal impact of health research. Strengthening institutional evaluation and reward systems to recognize KT-related activities may therefore encourage greater engagement in KT and ultimately enhance the translation of research evidence into policy and practice.

## Data availability

The data are available from the corresponding author upon request.

